# Identification of novel *Plasmodium vivax* proteins associated with protection against clinical malaria

**DOI:** 10.1101/2022.11.06.22282009

**Authors:** Ramin Mazhari, Eizo Takashima, Rhea J Longley, Shazia Ruybal-Pesantez, Michael T White, Bernard N Kanoi, Hikaru Nagaoka, Benson Kiniboro, Peter Siba, Takafumi Tsuboi, Ivo Mueller

## Abstract

As progress towards malaria elimination continues, the challenge posed by the parasite species *Plasmodium vivax* has become more evident. In many regions co-endemic for *P. vivax* and *Plasmodium falciparum*, as transmission has declined the proportion of cases due to *P. vivax* has increased. Novel tools that directly target *P. vivax* are thus warranted for accelerated elimination. There is currently no advanced vaccine for *P. vivax* and only a limited number of potential candidates in the pipeline. In this study we aimed to identify promising *P. vivax* proteins that could be used as part of a subunit vaccination approach. We screened 342 *P. vivax* protein constructs for their ability to induce IgG antibody responses associated with protection from clinical disease in a cohort of children from Papua New Guinea. This approach has previously been used to successfully identify novel candidates. We were able to confirm previous results from our laboratory identifying the proteins reticulocyte binding protein 2b and StAR-related lipid transfer protein, as well as at least four novel candidates with similar levels of predicted protective efficacy. Assessment of these *P. vivax* proteins in further studies to confirm their potential and identify functional mechanisms of protection against clinical disease are warranted.

## 1 Introduction

Malaria, an infectious disease caused by the parasite *Plasmodium*, remains a major global health problem. There have been substantial reductions in the burden of this disease throughout many endemic regions over the past two decades; however, in 2021, the World Health Organisation (WHO) reported that progress towards elimination had been disrupted due to the COVID-19 pandemic, with an estimated 14 million more cases in 2020 compared to 2019 (WHO, 2021b). Two species of *Plasmodium* parasites are responsible for most cases and deaths in humans: *P. falciparum* and *P. vivax*. Outside of sub-Saharan Africa, *P. vivax* is the most geographically widespread species and is quickly becoming responsible for most cases. This is evidenced by a marked shift in the epidemiology of malaria in co-endemic regions with decreasing transmission; in the Americas and Asia-Pacific the number of cases due to *P. falciparum* has steadily declined however *P. vivax* has persisted (Price et al., 2020). This highlights the urgent need for specific tools to target *P. vivax*, as routine clinical case management and vector control are not having the same effects as they did for *P. falciparum*. Failure of these preventative and control methods for *P. vivax* is likely due to several distinct biological features of this species, including an arrested stage in the liver that results in relapsing infections (the hidden hypnozoite reservoir) and earlier production of the sexual stages (gametocytes) that are required for onward transmission. There is also a large burden of asymptomatic (often low-density) *P. vivax* infections in low-transmission regions that escape routine detection (Harris et al., 2010;Waltmann et al., 2015;Nguitragool et al., 2017;Almeida et al., 2018;Sattabongkot et al., 2018).

The only successful method for eradicating any infectious disease affecting humans has been through vaccination. The first vaccine for malaria, RTS,S, has been endorsed by the WHO for broad use in children living in regions with moderate to high *P. falciparum* malaria transmission (WHO, 2021a). RTS,S targets a protein at the pre-erythrocytic stage of infection and is specific for *P. falciparum*. Unfortunately, for *P. vivax*, pre-clinical and clinical vaccine candidates are still lacking, with no *P. vivax* vaccine having progressed to phase II clinical trials in endemic regions (Draper et al., 2018;White and Chitnis, 2022). A key challenge for the development of a *P. vivax* vaccine has been the prioritisation of promising candidates. A subunit vaccine based on one or more *P. vivax* protein(s) is the most feasible for development (versus, for example, a whole parasite vaccine), however, more than 5000 proteins are expressed throughout the parasite’s lifecycle. Due to limitations in studying the biology of *P. vivax* (including the lack of a continuous culture system and accessible animal models) most progress in identifying potential candidate proteins for a vaccine has been through studies of naturally acquired immunity in endemic populations (Chia et al., 2014).

Evidence for naturally acquired immunity against *P. vivax* clinical disease has been gained through epidemiological observations, where studies have shown a decrease in morbidity with age or with successive exposure (reviewed in (Longley et al., 2016b)). Interestingly, the acquisition of immunity against clinical disease appears to be faster for *P. vivax* than *P. falciparum*, possibly due to the increased force of genetically distinct blood-stage infections seen for *P. vivax* (Koepfli et al., 2013) or other currently unknown mechanisms. Naturally acquired immunity is reliant on the generation of *Plasmodium*-specific antibodies (Cohen et al., 1961;Sabchareon et al., 1991), which perform various effector functions such as inhibition of invasion of red blood cells, neutralisation, opsonisation, and antibody-dependent cellular inhibition. Antibody responses following *P. vivax* infections can be long-lived (Wipasa et al., 2010;Longley et al., 2016a), which is a promising sign for vaccine development. However, in natural infection antibodies are induced to a broad array of *P. vivax* proteins (Finney et al., 2014), and the specific targets of a protective response have remained largely elusive.

We previously conducted a screen of IgG antibody responses against 38 *P. vivax* proteins in a cohort of children from Papua New Guinea (PNG) (Franca et al., 2017). High antibody levels to several *P. vivax* proteins, including novel targets, were strongly associated with reduced risk of clinical *P. vivax* infections. In the current study we used the same cohort to screen a much larger panel of *P. vivax* proteins (>300), with the aim of validating previous targets and identifying further novel candidates.

## 2 Materials and Methods

### 2.1 Study design overview

The goal of this study was to both validate existing and identify novel *P. vivax* proteins that are associated with protection against clinical disease in a population naturally exposed to infection. We therefore utilised plasma samples from a well-described longitudinal cohort study of Papua New Guinean children (Lin et al., 2010) and measured total IgG antibody responses in these children against 342 *P. vivax* protein constructs. We used statistical methods to assess the association of IgG antibody responses with various epidemiological factors and with protection against clinical disease.

### 2.2 Study samples: longitudinal cohort study in Papua New Guinea

Samples were obtained from a previously described longitudinal cohort study in children from PNG (Lin et al., 2010). Briefly, 264 children aged 1-3 years living in an area near Maprik, East Sepik Province, were enrolled from March to September 2006. Children were followed for 16 months with active monitoring of morbidity every two weeks. Finger-prick blood samples were collected every 8 weeks for molecular detection of *Plasmodium* spp. by PCR. All PCR-positive *P. vivax* infections were genotyped using the molecular markers *msp1*F3 and MS16. This allowed the incidence of genetically distinct blood-stage infections to be determined per child. This surrogate measure of individual differences in exposure is denoted as the molecular force of blood-stage infection (molFOB) (Koepfli et al., 2013). Plasma samples for the current study were used from 183 children from the first enrolment in March 2006 (Lin et al., 2010) who completed follow-up (see Figure 1). Clinical *P. vivax* episodes were defined as fever ≥37.5°C or reported history of fever within the last 48 hours with concurrent *P. vivax* parasitaemia >500 parasites/μl.

**Figure 1:**
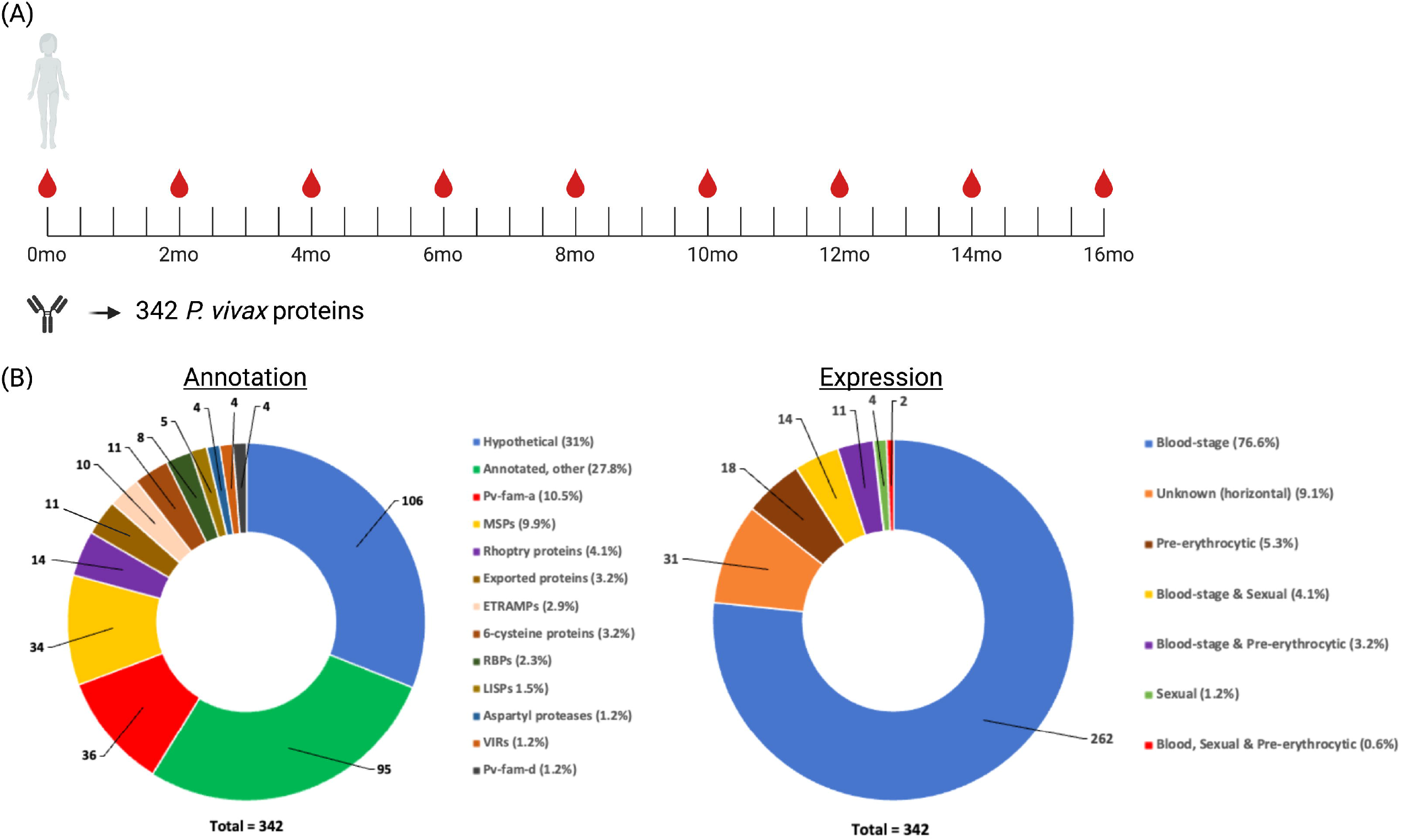
Overview of study design. **(A)** IgG antibodies were measured in 183 children at the first time-point of the longitudinal study. PCR data was available every 8 weeks throughout the 16-month long study, in addition to morbidity data that was obtained at every visit (every 2 weeks). **(B)** IgG antibodies were measured against 342 *P. vivax* proteins with the bottom plots providing a summary of the annotation and expression stages covered. Created with BioRender.com.

### 2.3 Ethical approval

Written informed consent was obtained from the parents or guardians of all children prior to enrolment. The study was approved by the Medical Research and Advisory Committee of the Ministry of Health in PNG (MRAC 05.19), and samples for use in this study by the Walter and Eliza Hall Institute of Medical Research Human Research Ethics Committee (07/07).

### 2.4 Antigen selection and production

A large panel of 342 *P. vivax* protein constructs were used in this study. This included *P. vivax* proteins selected due to the presence of immunogenic *P. falciparum* orthologs, or predicted signal peptides and/or transmembrane domains (as these factors increase the likelihood that the protein is exposed to the human immune response). These proteins are the same as in our recently published panel (Longley et al., 2020). For large proteins, more than one fragment was expressed and assessed for reactivity. In total, 303 unique proteins were screened. Key features of the protein are shown in Figure 1.

The proteins were expressed using the wheat germ cell-free protein expression system (CellFree Sciences, Matsuyama, Japan) as previously described (Longley et al., 2017b;Longley et al., 2020), noting that these proteins were not purified, and that the translation mixture was directly added into the antibody-screening assay as described below. All proteins were single-biotinylated.

### 2.5 Total IgG antibody measurements

Total IgG antibody measurements were made using the AlphaScreen assay following the manufacturer’s instructions (PerkinElmer Life and Analytical Sciences, Boston, MA) and as previously described (Longley et al., 2017b). Briefly, reactions were carried out in 384-well microtiter plates at 26 ºC using a JANUS Automated Workstation (PerkinElmer). 0.1µl of the translation mixture containing a biotinylated recombinant *P. vivax* protein was diluted 50-fold, mixed with 10µl of 4000-fold diluted plasma and incubated for 30 minutes to form an antigen-antibody complex. Next, streptavidin-coated donor-beads and acceptor-beads (PerkinElmer) conjugated with protein G were added and incubated for 1 hour in the dark to allow the donor and acceptor-beads to optimally bind to biotin and human IgG, respectively. Illumination of the formed complex resulted in a luminescence signal emitted at 620 nm, which was detected using an EnVision plate reader (PerkinElmer) with the result expressed as AlphaScreen counts. Each assay plate contained a standard curve of biotinylated rabbit IgG (at 0,6.25,12.5, 25, 50, 100, 200 and 400pM), enabling standardisation between plates using a 5-paramater logistic standard curve. Each AlphaScreen count was converted to an Arbitrary Unit between 0-400. Samples were run in singlicate only due to the limited plasma volume available.

We defined a seropositivity cut-off based upon the limit of detection of the assay as per our previous analysis using this panel of proteins and antibody detection method (Longley et al., 2017b). The sero-positivity cut-off was therefore set as half the lowest non-negative value, calculated on a per antigen basis. Seroreactivity was then defined as proteins where more than 10% of individuals had antibody levels above the sero-positivity cut-off, to enable direct comparison with our previously published datasets.

### 2.6 Statistical analysis

Statistical analysis was performed using R version 4.1.1. Arbitrary Units were log-10 transformed prior to statistical analysis. Spearman’s rank correlation test was used to associate IgG levels with age and molecular force of infection (molFOB), with a Bonferroni adjustment for multiple comparisons. A two-sided unpaired t-test was used to compare mean IgG levels between infected and uninfected children for each protein.

Two methods were used to assess for potential associations between total IgG responses to individual *P. vivax* proteins and protection against clinical episodes of *P. vivax:* (i) regression-based analysis (a conventional statistical method) and (ii) random forests (a machine-learning method). For the regression-based unadjusted analysis, we first implemented a logistic regression without adjusting for confounders to compare antibody levels measured at time point 0 months, with the presence or absence of an episode of clinical *P. vivax* within each of the eight 2-month intervals (see Figure 1). We then implemented an adjusted analysis using a generalised estimating equation (GEE) model as previously described (Franca et al., 2017), to account for within-individual correlation using an exchangeable structure. The model was further adjusted for individual differences in exposure (molFOB), village of residence, age, seasonality, and insecticide-treated net use. In a second approach, a Random Forests algorithm was used to identify features that were predictive of the incidence of *P. vivax* clinical cases. The tested feature included all antibody measurements and the epidemiological covariates defined above. The contribution of each feature to prediction performance was ranked using two metrics: mean decrease in accuracy and mean decrease in the Gini coefficient.

## 3 Results

### 3.1 Seroreactivity and epidemiological trends of the IgG response against the panel of 342 *P. vivax* proteins

IgG antibody responses were screened against a large panel of 342 *P. vivax* proteins in the Papua New Guinean children. We expected that most proteins would be seroreactive, given our previous results in a similar region of PNG in older children (5-10 years of age) (Longley et al., 2017b). In the current study, using a seropositivity cut-off of half the lowest non-negative value for each protein, more than 50% of children had detectable antibody responses to the entire panel of 342 *P. vivax* proteins (Supplementary Figure 1A). On an individual level, each child had an IgG response above background to between 7-342 *P. vivax* proteins. There were only 4 children seroreactive to <50% of the protein panel, with most children having detectable IgG responses to a large majority of the proteins tested (Supplementary Figure 1B).

Based on prior research it is expected that IgG antibodies to *P. vivax* proteins increase with age (Longley et al., 2017a), partially as age is a proxy for cumulative exposure. Despite the small (and young) age range of the children in the current study (1-3 years), we expected to detect an association with age for at least some of the *P. vivax* proteins based on our prior analyses in this cohort (Franca et al., 2017). However, no significant associations between IgG antibody level and age to any *P. vivax* antigen were observed (Spearman’s rank correlation adjusted p>0.05 for all antigens after Bonferroni correction for multiple comparisons). Given the young age of the children we also assessed associations with the molecular force of blood-stage infection (molFOB), a surrogate measure of exposure during follow-up (Franca et al., 2017). Individually, there were no statistically significant associations between life-time exposure and IgG level to any of the 342 proteins.

Despite no trend towards significant associations between IgG antibody levels and exposure (using either of age or molFOB), we did observe significant associations with current infection status (another epidemiological variable known to boost IgG levels (Longley et al., 2017a)). At the time of enrolment, when total IgG responses were measured, 91 (49.7%) children were currently infected with *P. vivax* (as determined by PCR) and we observed higher IgG levels to most proteins (n=339/342) in children with current *P. vivax* infections compared to those uninfected in this study (two-sided unpaired t-test with Bonferroni correction, adjusted p=<0.011).

### 3.2 IgG levels and prospective risk of clinical *P. vivax* malaria

Over the 16 months of follow-up, the subset of children included in this study experienced an incidence rate of 1.11 clinical episodes/year (342 episodes in 306.8 follow-up years), as defined in the methods. We applied a univariate analysis to assess associations of IgG to each antigen with protection from clinical *P. vivax* episodes. Children were split into terciles based on the measured IgG antibody response at the first time point (low, medium, or high). After adjustment for molFOB, village, age, seasonality, and insecticide-treated bed net usage using a GEE model, high IgG antibody levels to most of the 342 *P. vivax* proteins associated with reduced risk of clinical malaria. However, the high degree of correlation between all *P. vivax* proteins (Supplementary Figure 2) suggests that many biomarkers may not be causally associated with protection. The data is presented as the potential protective efficacy (PPE), which is calculated by comparing the incidence of clinical *P. vivax* malaria in the high tercile compared to the low tercile (antibody) responders (Figure 2).

**Figure 2:**
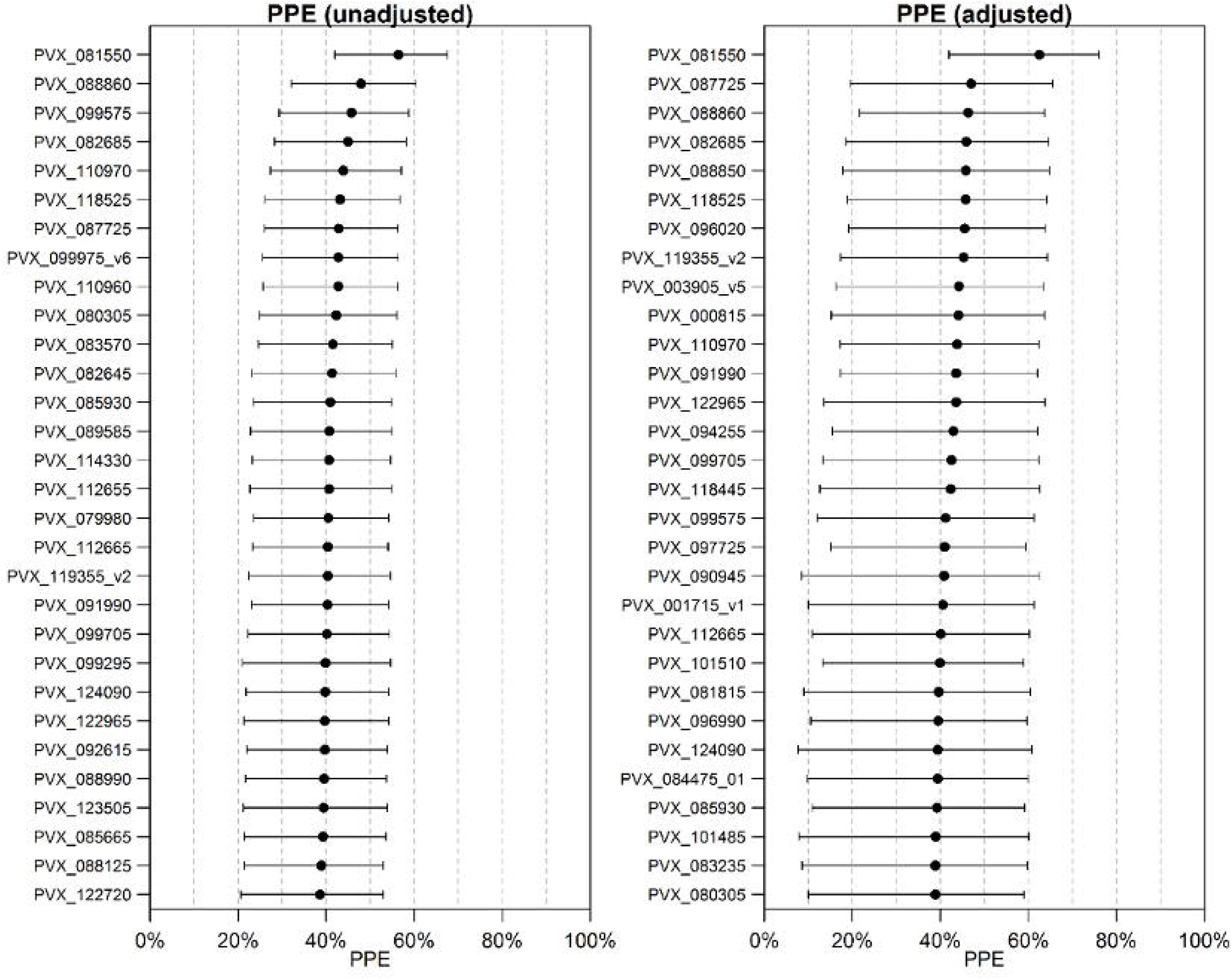
Estimated potential protective efficacy (PPE) for the top 30 *P. vivax* proteins. A) unadjusted analysis using logistic regression and B) adjusted analysis using a GEE model.

Associations with protection from clinical *P. vivax* infections were also assessed using a Random Forests algorithm with the data on antibody levels incorporated as continuous quantitative measurements. In this analysis the covariates are included into the classifier. The results are shown as variable importance plots (Figure 3). Notably, the molecular force of infection (pvfoinew) is the strongest predictor for both ways of quantifying contribution to classification performance. The top-ranked antigen (PVX_003555_1) in the variable importance plot is associated with increased risk of *P. vivax* clinical episodes rather than protection from *P. vivax* clinical episodes.

**Figure 3:**
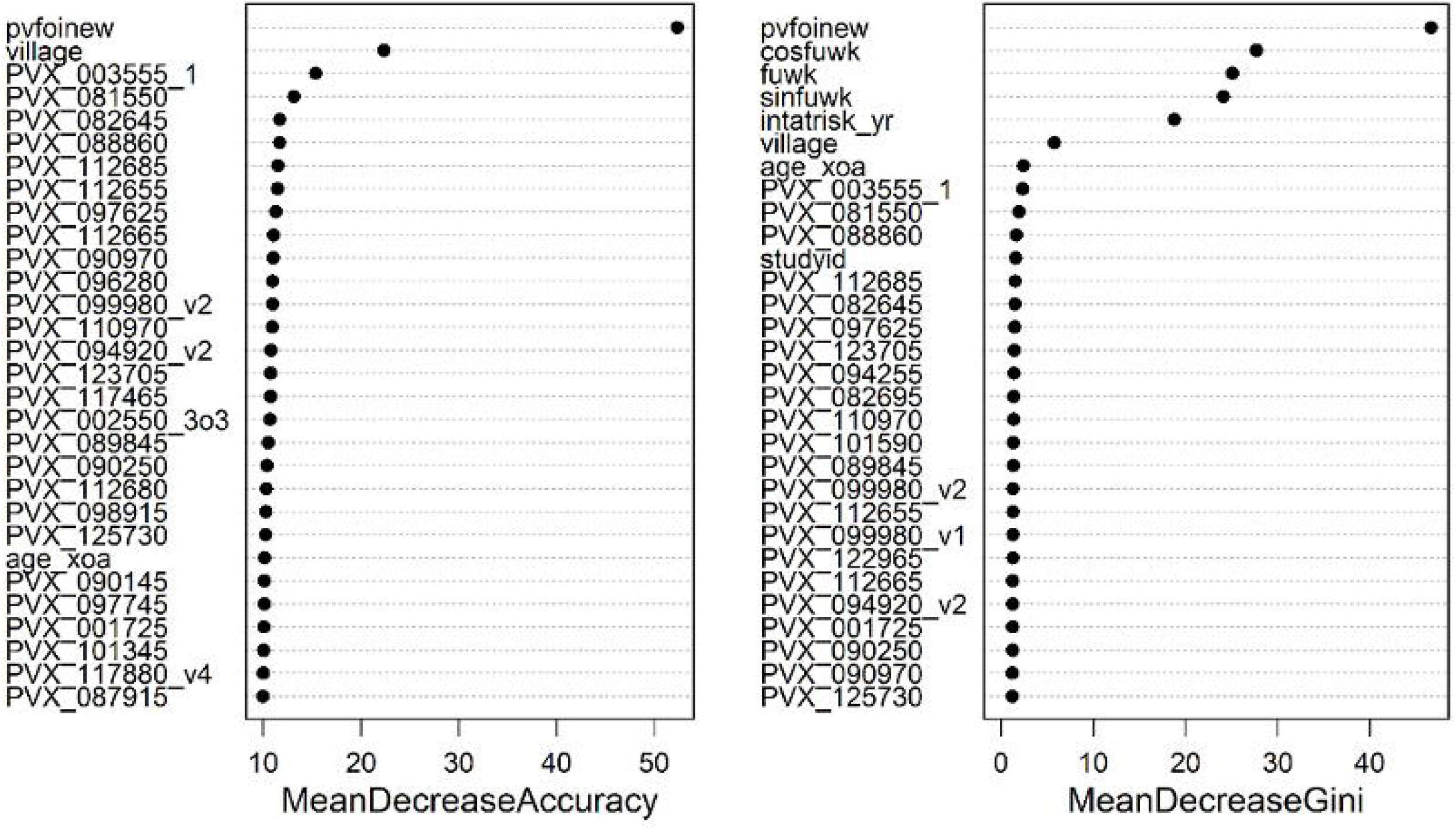
Variable importance plots from the Random Forests algorithm. Two measures of importance are given for each variable. The mean decrease in accuracy is based on how much the accuracy decreases when the variable is excluded. The second measure is based on the decrease of Gini impurity when a variable is chosen to split a node in a decision tree of the random forest. Higher ranking variables indicate a greater contribution to classification performance, i.e. between children with and without clinical *P. vivax* infections.

IgG antibody levels to the 342 *P. vivax* proteins were correlated to various extents (R=0.18 to 0.99, 82% with R >0.70) (Supplementary Figure 2). Therefore, to assist ranking the 342 *P. vivax* proteins based on their association with protection against clinical *P. vivax* episodes, rather than looking only at single protein associations we also looked at combinations of two or three proteins using the GEE model. Even using combinations of up to only three proteins, with 342 *P. vivax* constructs this equates to 342*341*340/(1*2*3) = 6,667,233 unique combinations. We used the PPE from single, two or three antigen combinations in adjusted regression models to rank the 342 *P. vivax* proteins. Table 1 shows the Top 20 ranked proteins for each method.

**Table 1:** Comparison of antigen ranking using single proteins versus combinations of two or three. Those in colour were ranked highly through multiple methods (regression and random forests, in one or multiple antigen combinations, or by both adjusted and unadjusted analyses).

| Rank | 1 antigen regression |  | 2 antigen regression |  | 3 antigen regression |  | random forests |  |
| --- | --- | --- | --- | --- | --- | --- | --- | --- |
|  | *Unadjusted* | *Adjusted* | *Unadjusted* | *Adjusted* | *Unadjusted* | *Adjusted* | *Accuracy* | *Gini* |
| 1 | PVX_081550 | PVX_081550 | PVX_081550 | PVX_081550 | PVX_081550 | PVX_081550 | PVX_081550 | PVX_081550 |
| 2 | PVX_088860 | PVX_087725 | PVX_088860 | PVX_094255 | PVX_110960 | PVX_094255 | PVX_082645 | PVX_088860 |
| 3 | PVX_099575 | PVX_088860 | PVX_110960 | PVX_097725 | PVX_088860 | PVX_097725 | PVX_088860 | PVX_112685 |
| 4 | PVX_082685 | PVX_082685 | PVX_097725 | PVX_092990_v1 | PVX_099295 | PVX_092990_v1 | PVX_112685 | PVX_082645 |
| 5 | PVX_110970 | PVX_088850 | PVX_092990_v1 | PVX_082685 | PVX_097725 | PVX_082680 | PVX_112655 | PVX_097625 |
| 6 | PVX_118525 | PVX_118525 | PVX_110970 | PVX_088860 | PVX_092990_v1 | PVX_110960 | PVX_097625 | PVX_123705 |
| 7 | PVX_087725 | PVX_096020 | PVX_099295 | PVX_110970 | PVX_112655 | PVX_082685 | PVX_112665 | PVX_094255 |
| 8 | PVX_099975_v6 | PVX_119355_v2 | PVX_099575 | PVX_082680 | PVX_099575 | PVX_115355 | PVX_090970 | PVX_082695 |
| 9 | PVX_110960 | PVX_003905_v5 | PVX_082685 | PVX_087725 | PVX_110970 | PVX_099980_v3 | PVX_096280 | PVX_110970 |
| 10 | PVX_080305 | PVX_000815 | PVX_112655 | PVX_099980_v3 | PVX_082685 | PVX_088860 | PVX_099980_v2 | PVX_101590 |
| 11 | PVX_083570 | PVX_110970 | PVX_080305 | PVX_118525 | PVX_089585 | PVX_122965 | PVX_110970 | PVX_089845 |
| 12 | PVX_082645 | PVX_091990 | PVX_082645 | PVX_110960 | PVX_080305 | PVX_110970 | PVX_094920_v2 | PVX_099980_v2 |
| 13 | PVX_085930 | PVX_122965 | PVX_094255 | PVX_122965 | PVX_094255 | PVX_087725 | PVX_123705 | PVX_112655 |
| 14 | PVX_089585 | PVX_094255 | PVX_089585 | PVX_115355 | PVX_123505 | PVX_080305 | PVX_117465 | PVX_099980_v1 |
| 15 | PVX_114330 | PVX_099705 | PVX_123505 | PVX_088850 | PVX_079980 | PVX_118525 | PVX_002550_3o3 | PVX_122965 |
| 16 | PVX_112655 | PVX_118445 | PVX_115355 | PVX_096020 | PVX_115355 | PVX_113965_v1 | PVX_089845 | PVX_112665 |
| 17 | PVX_079980 | PVX_099575 | PVX_079980 | PVX_080305 | PVX_082645 | PVX_087725 | PVX_090250 | PVX_094920_v2 |
| 18 | PVX_112665 | PVX_097725 | PVX_087725 | PVX_099295 | PVX_000975_v1 | PVX_080305 | PVX_112680 | PVX_001725 |
| 19 | PVX_119355_v2 | PVX_090945 | PVX_099975_v6 | PVX_119355_v2 | PVX_088990 | PVX_118525 | PVX_098915 | PVX_090250 |
| 20 | PVX_091990 | PVX_001715_v1 | PVX_088990 | PVX_099705 | PVX_121920 | PVX_113965_v1 |  |  |

### 3.3 Potential protective efficacy of IgG antibodies to combinations of *P. vivax* proteins

We further assessed how the PPE may improve with use of IgG antibody responses to more than 3 *P. vivax* antigens (Figure 4). Antigens were included in a stepwise manner by addition of the “next best” antigen (note that this does not guarantee the optimal combination). We compared the PPE between our current data and our previously published dataset in the same PNG child cohort (Franca et al., 2017), where IgG antibody responses were measured to 40 purified *P. vivax* proteins in a multiplexed Luminex assay. Using the same methods, we see that for n<15 Luminex performs better, despite the substantially smaller panel of antigens to select for. For n≥15, Alpha Screen performs better, with PPE eventually approaching 100%.

**Figure 4:**
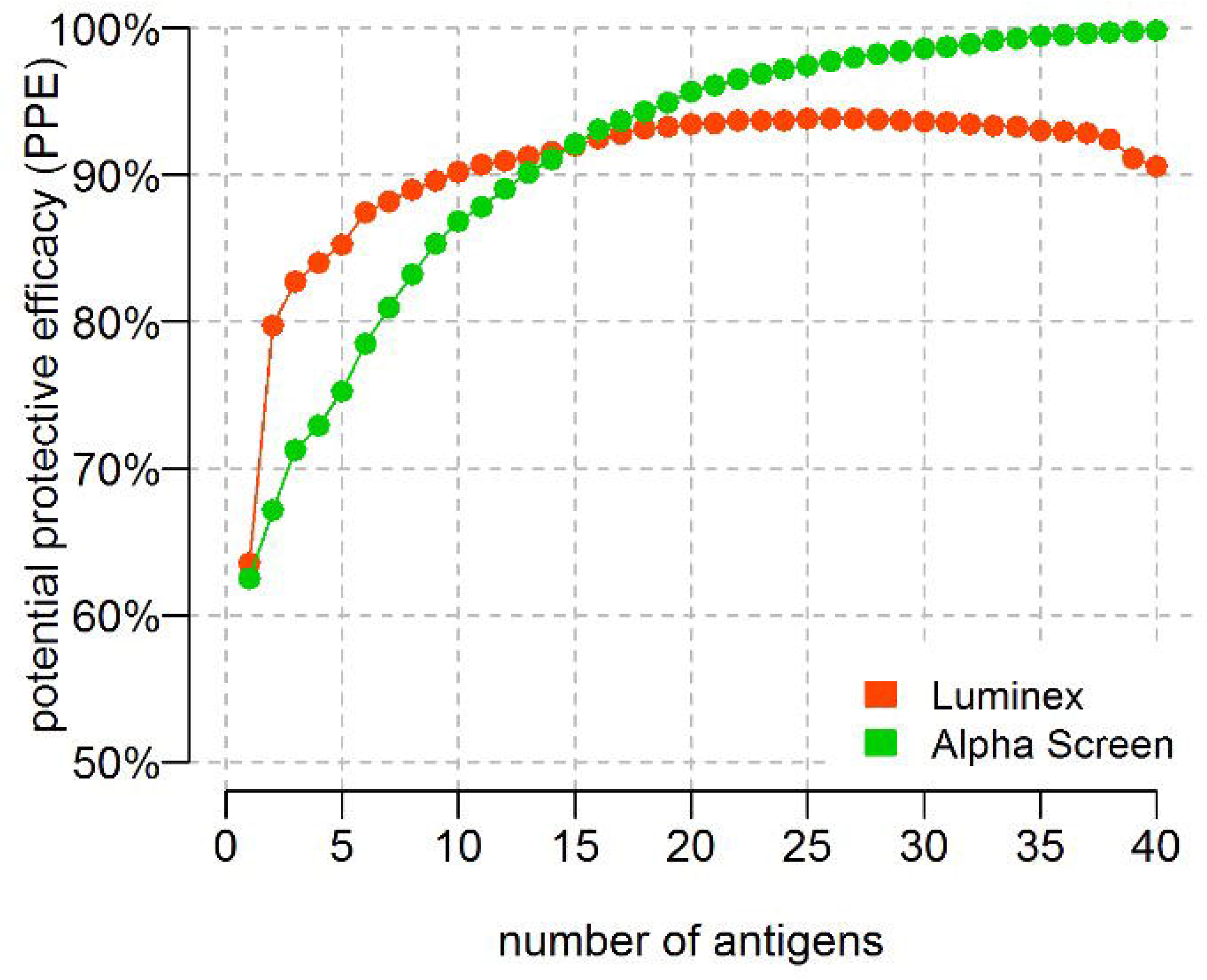
PPE with increasing numbers of *P. vivax* antigens. PPE with increasing numbers of antibody responses against *P. vivax* antigens included in a stepwise manner. Green: AlphaScreen data and Red: Luminex data. Note the antigens used on each platform were not the same.

## 4 Discussion

Persistence of *P. vivax* transmission despite control efforts is a major challenge for malaria elimination in South America and Asia. In addition, whilst there has been progress in the development of a *P. falciparum* vaccine, there is a lack of *P. vivax* vaccine candidates at all stages of the vaccine development pipeline (Reyes-Sandoval, 2021). We have recently used a well-characterised longitudinal cohort study of children in *P. vivax*-endemic PNG to identify *P. vivax* antigens that induce IgG antibodies associated with subsequent protection from clinical disease (Franca et al., 2017). In the current study we aimed to screen a much larger panel of *P. vivax* antigens (>300) in the same PNG child cohort to a) identify novel candidates for further assessment and b) potentially validate some of the candidates we previously identified. A similar approach has also been successfully pursued for *P. falciparum* (Kanoi et al., 2017). We overall observed a lack of association of IgG levels to the 342 *P. vivax* antigens with age and a measure of lifetime exposure, whilst IgG levels were significantly increased in children with concurrent *P. vivax* infections at the time of antibody measurement. Most *P. vivax* antigens were individually associated with protection from clinical disease, after adjustment for potential confounders. However, when a single biomarker appears associated with protection, this can often be due to correlation with other biomarkers, and it does not necessarily imply causality.

We utilised two methods for assessing the association between IgG levels at the start of the cohort with prospective risk of symptomatic *P. vivax* infections (a regression model and a Random Forests algorithm). Across both methods (and unadjusted and adjusted analyses), there were a number of *P. vivax* proteins that were highly ranked, including: PVX_081550 (StAR-related lipid transfer protein), PVX_088860 (sporozoite invasion-associated protein 2, SIAP2), PVX_094255 (reticulocyte binding protein 2b, RBP2b), PVX_087725 (hypothetical), PVX_110960 (hypothetical), PVX_082645 (merozoite surface protein 7, MSP7), PVX_082685 (MSP7), PVX_097725 (merozoite surface protein 3, MSP3), PVX_092990 (tryptophan-rich antigen (Pv-fam-a) and PVX_099295 (hypothetical). These 10 *P. vivax* antigens cover the top 4 from each method. A number of these constitute novel *P. vivax* antigens associated with protection and thus warrant further studies to validate these findings and determine the functional relevance of the antigens (including for the hypothetical proteins and the Pv-fam-a protein, which are relatively understudied). In addition, the top 10 listed includes several well-characterised *P. vivax* antigens with known roles in invasion, including RBP2b and the MSPs. RBP2b mediates binding of *P. vivax* parasites to reticulocytes through the Transferrin receptor 1 (Gruszczyk et al., 2018), and antibodies against RBP2b have previously been associated with protection from clinical disease in the same PNG child cohort (Franca et al., 2016) and in additional longitudinal studies in Brazil and Thailand (He et al., 2019). MSP7 and MSP3 are both multi-gene families, and whilst the exact proteins identified have not previously been associated with protection other family members have, most notably MSP3a (Stanisic et al., 2013;Franca et al., 2017). PVX_082645 (referred to as MSP7A), has been identified as the most abundant and immunogenic of the MSP7 family members (Cheng et al., 2019)). SIAP2 is the only antigen of the 10 listed that is known to be expressed primarily at the sporozoite stage (Roth et al., 2018), but to our knowledge has not previously been assessed for an association with clinical protection. The StAR-related lipid transfer protein was one of our top hits in our prior study (Franca et al., 2017).

In addition to successfully identifying novel candidates associated with clinical protection we also aimed to validate some of our prior targets associated with protection. There were a few differences between our earlier study and the current, including the panel of *P. vivax* antigens (40 vs 342), the expression and purification methods (notably our earlier study used purified proteins whilst the current used crude proteins), and the subset of children (225 in the prior study vs 183 in the current). Furthermore, not all 40 *P. vivax* antigens in the earlier study were included in our panel of 342. Of those that were included in both studies, two were amongst our top 10 as previously mentioned (RBP2b and the StAR-related lipid transfer protein). These were both within the top 5 antigens associated with protection in our earlier work and thus our current study validates their identification as promising *P. vivax* vaccine candidates. The remaining antigens within the top 5 individually associated with protection from clinical *P. vivax* episodes in our prior study were the erythrocyte binding protein (KMZ83376.1) (EBP), cysteine-rich protective antigen (CyRPA) (PVX_090240) and MSP3a (PVX_097720). The latter two proteins, CyRPA and MSP3a, were included in our current screen but were not identified by any method as within the top 20 *P. vivax* proteins associated with protection from clinical disease. This could feasibly be due to the different protein constructs and expression and purification systems used. Interestingly, when looking at the maximum potential protective efficacy reached when combining antibody responses to multiple *P. vivax* antigens, our prior study indicated a plateau once 20 antigens were used. When using our current dataset with an expanded number of *P. vivax* proteins we eventually approach 100% potential protective efficacy with a combination of antibody responses to 40 *P. vivax* antigens, however it is likely that this is due to overfitting of the data rather than a true biological effect.

Overall, the 342 *P. vivax* proteins were seroreactive in these young PNG children. However, a limitation of our study was the absence of any plasma samples from non-malaria endemic areas assessed in the same platform. Thus, the seroreactivity cut-off is reflective of responses above assay background rather than a true seropositivity cut-off. This issue is avoided in the analyses assessing associations with protection by splitting the IgG antibody data in terciles. The breadth of the response was large amongst these PNG children with most reactive to at least half of the *P. vivax* proteins assessed, which fits with prior estimates postulated in the literature (Finney et al., 2014;Longley et al., 2017b). There were also strong pairwise correlations in antibody levels between many proteins. We have observed this phenomenon previously in other studies of naturally acquired antibody responses against *P. vivax* proteins (Franca et al., 2017;Longley et al., 2020), and this could be related to co-acquisition of antibodies or cross-reactivity between targets. Surprisingly, we observed no association between antibody levels to most of the *P. vivax* proteins with age. Whilst this may be due to the young and limited age range of the children (1-3 years), we had previously observed clear positive associations with age in our earlier study in this same cohort (Franca et al., 2017). Thus, the absence of associations with age may be due to the difference in the technical platforms used to measure antibody responses. In addition, we also failed to observe associations between most antibody responses and lifetime exposure (using molFOB as a surrogate). However, we did observe a significant positive association between concurrent *P. vivax* infections and the antibody response to almost all 342 *P. vivax* proteins as expected.

The combination of the wheat germ cell-free protein expression system and the AlphaScreen platform utilised in this study is a powerful approach to screen antibody responses to large panels of *P. vivax* proteins (Kanoi et al., 2021). Here, we identify novel antigens that induce IgG antibody responses associated with protection from clinical *P. vivax* infections (including the hypothetical proteins PVX_087725, PVX_110960 and PVX_099295), as well as validate previously identified targets (RBP2b, StAR-related lipid transfer protein). These, in addition to the other top 20 *P. vivax* antigens identified, warrant further assessment using purified proteins and other antibody screening assays such as ELISA and Luminex. Furthermore, measurement of IgG magnitude alone does not infer whether the antibody responses detected are markers of immunity or functionally relevant in providing protection. Additional analyses can be undertaken to measure markers of functional antibody responses, such as the ability to fix complement or bind Fcy receptors, and subsequently lower throughput assays to directly measure functional activity such as invasion inhibition assays (Opi et al., 2021). Together these can provide novel insights into *P. vivax* antigens of interest for immune-based interventions such as vaccines or monoclonal antibodies.

## Data Availability

Newly generated AlphaScreen data is provided as Supplementary Data 1. Epidemiological data required for associations with protection analyses is provided in Supplementary Data 2. All data and code for the seroreactivity and epidemiological analyses are available at: https://github.com/shaziaruybal/R03-alphascreen-analysis.

## 7 Author Contributions

RM, RJL, MTW, ET, TT and IM conceived the study. BK, PS and IM collected study samples. RM, BNK, HN performed experiments. RM, RJL, MTW, SR-P performed data analysis. All authors reviewed and approved the final version of the manuscript.

## 9 Acknowledgments

We gratefully acknowledge all individuals and their families participating in this study. We thank the large Papua New Guinea Team for conducting the field study. We thank Dr Connie Li-Wai-Suen for assisting in converting the raw AlphaScreen counts to arbitrary units based on the standard curve.

