## Supplementary Material (Figure S1, Figure S2) for "Identification of novel *Plasmodium vivax* proteins associated with protection against clinical malaria"

### 1 Supplementary Data

Supplementary Data 1: AlphaScreen data.

Supplementary Data 2: Epidemiological data.

### 2 Supplementary Figures and Tables

#### 2.1 Supplementary Figures

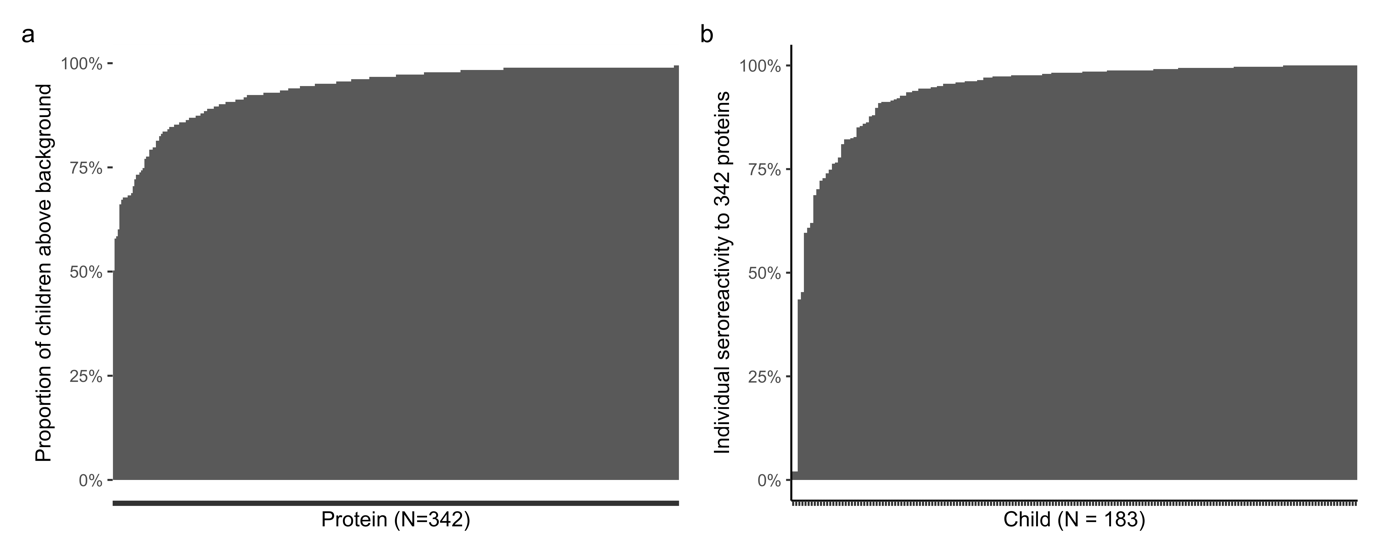

**Supplementary Figure 1.** **Seroreactivity of the 342 *P. vivax* proteins in Papua New Guinean children.** A) The proportion of children with IgG levels above the defined background (half the lowest non-negative value for each protein). All proteins had more than 50% of children above background. B) Variation in individual seroreactivity. Data shown is the proportion of the protein panel (n=342 proteins) each child is seroreactive against.

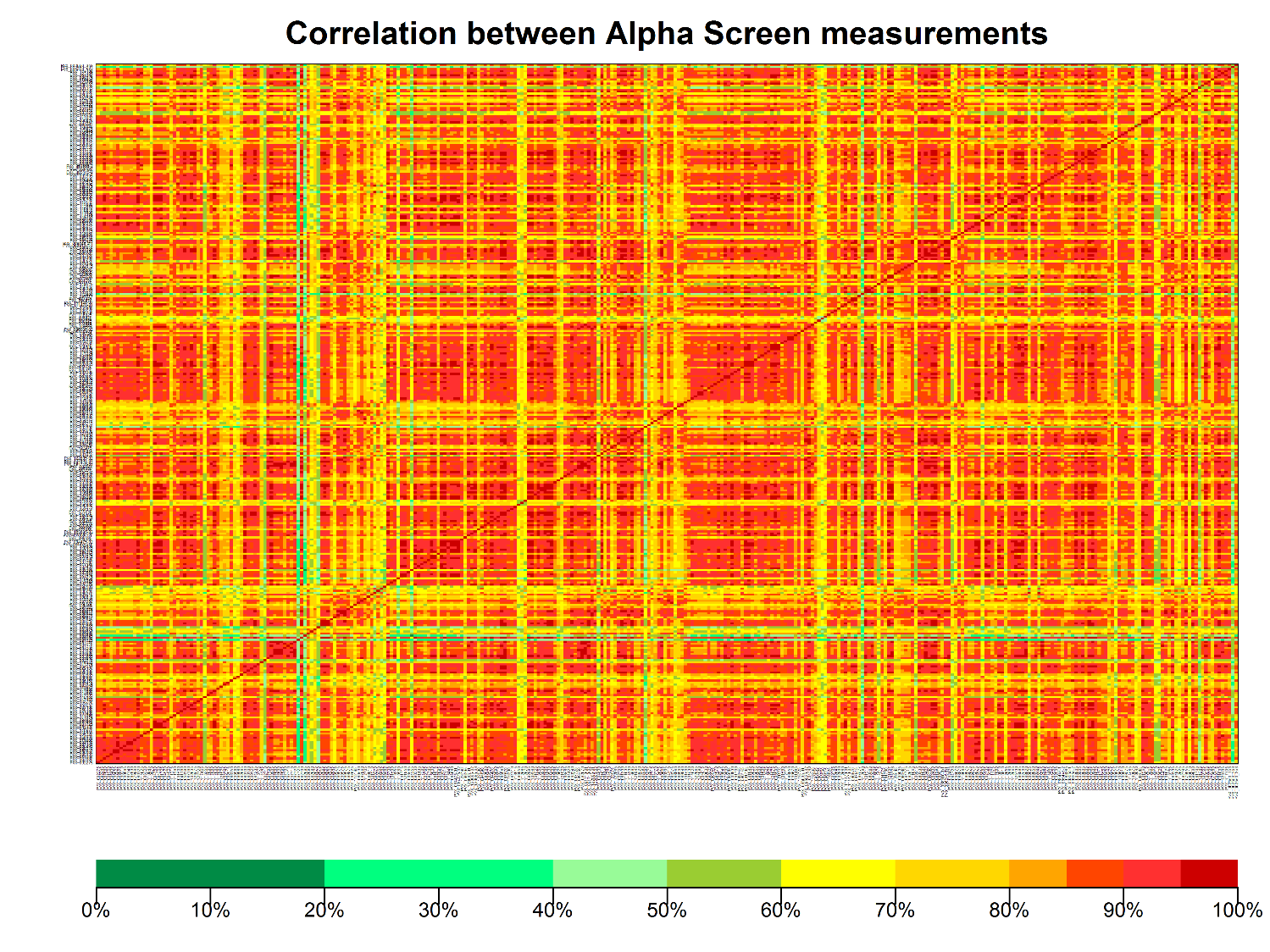

**Supplementary Figure 2. Correlation between IgG measurements to 342 *P. vivax* proteins.** Pairwise correlations of IgG Antibody Units between all 342 *P. vivax* antigens.
